# Motor Function and White Matter Connectivity in Children Treated with Therapeutic Hypothermia for Neonatal Encephalopathy

**DOI:** 10.1101/2021.04.30.21256369

**Authors:** Arthur P.C. Spencer, Jonathan C.W. Brooks, Naoki Masuda, Hollie Byrne, Richard Lee-Kelland, Sally Jary, Marianne Thoresen, Marc Goodfellow, Frances M. Cowan, Ela Chakkarapani

## Abstract

Therapeutic hypothermia reduces the incidence of severe motor disability, such as cerebral palsy, following neonatal hypoxic-ischemic encephalopathy. However, cooled children without cerebral palsy at school-age demonstrate motor deficits and altered white matter connectivity. In this study, we used diffusion-weighted imaging to investigate the relationship between white matter connectivity and motor performance, measured using the Movement Assessment Battery for Children-2, in school-age children treated with therapeutic hypothermia for neonatal hypoxic ischaemic encephalopathy at birth, who did not develop cerebral palsy (cases), and matched controls. Analysis of tract-level microstructure (33 cases, 36 controls) revealed correlations between total motor scores and fractional anisotropy, in cases but not controls, in the anterior thalamic radiation bilaterally, the inferior fronto-occipital fasciculus bilaterally and both the hippocampal and cingulate gyrus parts of the left cingulum. Analysis of structural brain networks (22 cases, 32 controls), in which edges were determined by probabilistic tractography and weighted by fractional anisotropy, revealed correlations between total motor scores and several whole-brain network metrics in cases but not controls. We then investigated edge-level association with motor function using the network-based statistic. This revealed subnetworks which exhibited group differences in the association between motor outcome and edge weights, for total motor scores as well as for balance and manual dexterity domain scores. All three of these subnetworks comprised numerous frontal lobe regions known to be associated with motor function, including the superior frontal gyrus and middle frontal gyrus. These findings demonstrate an association between impaired motor function and brain organisation in case children.

## Introduction

Neonatal hypoxic ischaemic encephalopathy (HIE), secondary to perinatal asphyxia, increases the risk of death and disability, including cerebral palsy (CP) (Marlow, 2005; O’Connor et al., 2017; Robertson et al., 1989). Even in the absence of CP, children who suffered HIE can develop motor impairments by school age (de Vries and Jongmans, 2010; van Kooij et al., 2010, 2008; van Schie et al., 2015). The treatment for NE, as recommended in the UK by the National Institute for Clinical Excellence (https://www.nice.org.uk/guidance/ipg347), is therapeutic hypothermia (TH), which involves cooling the infant’s core temperature to 33.5°C for 72 hours commencing as soon as possible after the asphyxia (Azzopardi et al., 2009; Rutherford et al., 2010). TH improves outcome compared to non-cooled children with HIE, reducing the chance of death, disability, and severe motor impairment including CP (Azzopardi et al., 2014; Jacobs et al., 2013; Jary et al., 2015). However, recent studies have shown that school-age children who received TH at birth for HIE, and did not develop CP, exhibited motor and cognitive impairments to a degree not found in typically developing controls (Jary et al., 2019; Lee-Kelland et al., 2020; Tonks et al., 2019). Additionally, motor impairment at school age was not predicted by motor performance assessed using Bayley Scales of Infant & Toddler development at 18 months of age (Jary et al., 2019). These studies suggest that, despite the success of TH in reducing the occurrence of severe disabilities, aspects of brain development may remain affected by HIE. In our previous work, we identified disrupted white matter connectivity in school-age children without CP, who were given TH for HIE, and demonstrated an association between these structural connectivity deficits and cognitive impairments (Spencer et al., 2021). It is not yet understood how brain structure relates to motor ability following TH; understanding this relationship will provide insight into damage mechanisms following HIE treated with TH which alter development, and may inform follow-up care and the development of new interventions.

White matter microstructure and its large-scale structural connectivity are associated with motor function (Englander et al., 2015; López-Vicente et al., 2021). As white matter connectivity undergoes substantial changes around birth (Dennis and Thompson, 2013a; Dubois et al., 2014), with microstructural alterations continuing into adolescence (Cascio et al., 2007; Hagmann et al., 2010; Lebel et al., 2008; Simmonds et al., 2014), insult or injury at birth (such as HIE) can have a lasting impact on brain organisation and functional outcome (Hüppi and Dubois, 2006). Diffusion-weighted imaging (DWI) allows non-invasive investigation of white matter organisation by measuring the diffusion of water molecules through brain tissue. This allows measurement of diffusion metrics such as fractional anisotropy (FA), which is affected by myelination and fibre density (Le Bihan and Johansen-Berg, 2012), offering clinically relevant characterisation of white matter microstructure (Assaf et al., 2019; Assaf and Pasternak, 2008; Dennis and Thompson, 2013b). This can be extended to the analysis of large-scale brain connectivity using a network neuroscience approach (Bassett and Sporns, 2017; Bullmore and Sporns, 2009; Fornito et al., 2013; Sporns et al., 2005). Structural brain networks, or connectomes, can be constructed by performing tractography in order to determine connections (edges) between brain regions (nodes), and weighting applied to edges according to diffusion properties of the white matter through which they track. Properties of brain networks can then be quantified using techniques from graph theory. This approach has been widely used to study typical and atypical brain development (Dennis and Thompson, 2013a; Hagmann et al., 2010; Morgan et al., 2018; Smyser et al., 2019).

In this study, we investigated the association between white matter connectivity and motor function in school-age children treated with TH for HIE at birth, who did not develop CP, compared to controls with no overt neurological problems, matched for age, sex and socio-economic status. We measured the correlation between motor outcome and tract-level microstructure using an age-specific atlas of white matter tracts (Spencer et al., 2020). We then constructed FA-weighted structural brain networks for each subject, using probabilistic tractography, and investigated the relationship between graph-theoretic network metrics and motor outcome. In order to further explore the relationship between brain organisation and motor function, we used the network-based statistic (NBS) (Zalesky et al., 2010) to determine subsets of connections (subnetworks) which exhibited group differences in the dependence of motor scores on edge weight.

## Materials and Methods

### Participants

Informed and written consent was obtained from the parents of participants, in accordance with the Declaration of Helsinki. Ethical approval was obtained from the North Bristol Research Ethics Committee and the Health Research Authority (REC ID: 15/SW/0148).

Case children were sequentially selected from the cohort of children who received TH between 2008 and 2011. These data are maintained by the Bristol Neonatal Neurosciences group at St Michael’s Hospital, Bristol, UK, under previous ethics approval (REC ID: 09/H0106/3). Eligibility criteria for the cases included: gestation at birth ≥36 weeks; received treatment with TH as standard clinical care based on TOBY trial eligibility criteria including signs of perinatal asphyxia and moderate to severe encephalopathy (confirmed by clinical examination and amplitude integrated electroencephalogram (Azzopardi et al., 2009)); cooling administered within six hours of birth and for at least 72 hours. Children were excluded if they had been found to have a metabolic or genetic disorder or if any major intracranial haemorrhage or structural brain abnormality could be seen on the neonatal MRI scan, if they were not native English speakers, or if they had any additional medical diagnosis other than HIE. All case children underwent neonatal MRI, which was qualitatively assessed, by an experienced perinatal neurologist (FC), for the presence and extent of brain injury. This was quantified, in the basal ganglia and thalami (scored 0-3), white matter (scored 0-3) and the posterior limb of internal capsule (scored 0-2), where a higher number indicates more severe injury (Rutherford et al., 2010; Skranes et al., 2017). A diagnosis of CP was ruled out at 2 years and at 6-8 years based on assessment of motor function using a standard clinical neurological examination including assessment of tone, motor function and deep tendon reflexes.

The control group consisted of children matched for age, sex and socio-economic status (Lee-Kelland et al., 2020), recruited via local schools in Bristol, UK and newsletters circulated at the University of Bristol. Exclusion criteria for controls were as follows: born before 36 weeks gestation; had any history of HIE or other medical problems of a neurological nature (confirmed using the same neurological clinical examination as for cases); were not native English speakers.

Socio-economic status was measured using the index of multiple deprivation as defined by the UK Government (www.gov.uk/government/statistics/english-indices-of-deprivation-2019) based on post code at birth. This is measured on a scale of 1–10, computed from 7 domains of deprivation including income, employment, education, health, crime, barriers to housing & services and living environment, indicating the decile within which the local area is ranked in the country, from most deprived to least deprived.

### Motor Assessment

Assessment of motor function was carried out using the Movement Assessment Battery for Children, Second Edition (MABC-2) (Henderson et al., 2007). We used MABC-2 as it has high test-retest reliability, content, construct and criterion validity and has evidence of predictive validity (Griffiths et al., 2018). This consists of eight raw test scores summarised into three subscales (aiming and catching, balance, and manual dexterity), each normalised according to a standardised sample with a mean (standard deviation) of 10 (3). The sum of all test scores is used to calculate the MABC-2 total score. MABC-2 total scores between the 6th and 15th centiles indicate a high risk of motor difficulty, and scores ≤5th centile indicate significant motor difficulty. Assessments were videoed and double-scored by a further assessor unaware of case status. Discrepancies between scores were agreed by consensus.

### MRI Acquisition

As previously reported (Spencer et al., 2021), images were acquired with a Siemens 3 tesla Magnetom Skyra MRI scanner at the Clinical Research and Imaging Centre (CRiCBristol), Bristol, UK. A child-friendly, detailed explanatory video was developed (EC and RLK) and sent to family at home before assessment day if they wanted and also shown and discussed with children on the day just prior to the scan. Additionally, experience of the typical sounds audible in the MRI scanner were provided. Children were placed supine within the 32-channel receive only head-coil by an experienced radiographer, and head movement was minimised with memory-foam padding. Children wore earplugs and were able to watch a film of their choice. A sagittal volumetric T1-weighted anatomical scan was acquired with the magnetisation-prepared rapid acquisition gradient echo (MPRAGE) sequence using the following parameters: echo time (TE) = 2.19 ms; inversion time (TI) = 800 ms; repetition time (TR) = 1500 ms; flip angle = 9°; field of view (FoV) 234 × 250 mm; 176 slices; 1.0 mm isotropic voxels. DWI data were acquired with a multiband echo-planar imaging (EPI) sequence, using the following parameters: TE = 70 ms; TR = 3150 ms; FoV 192 × 192 mm; 60 slices; 2.0 mm isotropic voxels, flip angle 90°, phase encoding in the anterior-posterior direction, in-plane acceleration factor = 2 (GRAPPA (Griswold et al., 2002)), through-plane multi-band factor = 2 (Moeller et al., 2010; Setsompop et al., 2012b, 2012a). Two sets of diffusion-weighted images, each with b = 1,000 s mm^-2^ in 60 diffusion directions and an additional eight interspersed b = 0 images, were acquired with a blip-up blip-down sequence, giving a total of 136 volumes.

### Pre-processing

T1-weighted images were denoised with the Advanced Normalization Tools DenoiseImage tool (http://github.com/ANTsX/ANTs) (Manjón et al., 2010). Brain tissue was extracted using either SPM8-VBM (http://fil.ion.ucl.ac.uk/spm) (Ashburner and Friston, 2005) or CAT12 (http://www.neuro.uni-jena.de/cat) (Gaser and Dahnke, 2016) depending on which gave better delineation of the brain surface for each subject. DWI data were corrected for eddy current induced distortions and subject movements using EDDY (Andersson and Sotiropoulos, 2016) and TOPUP (Andersson et al., 2003) from the FMRIB Software Library (FSL, http://fsl.fmrib.ox.ac.uk) (Smith et al., 2004).

### Quality Control

T1-weighted images were assessed visually and rejected if they had any severe movement artefacts. The structural pipeline described below was then applied to the remaining scans, followed by further visual inspection of the parcellation and tissue segmentation. Scans were rejected at this stage if any moderate artefacts had caused errors in the parcellation or segmentation. The quality of the DWI data was assessed using the EddyQC tool (Bastiani et al., 2019) from FSL, which gives metrics indicating the level of movement and eddy currents in each direction. Scans were rejected if the root-mean-square of these metrics was greater than one standard deviation above the mean for the whole cohort.

### Tract Microstructure

We assessed the relationship between MABC-2 total score and FA in the anterior thalamic radiation (ATR), cingulate gyrus part of the cingulum (CG), hippocampal part of the cingulum (CH), corticospinal tract (CST), forceps minor (Fminor), forceps major (Fmajor), inferior fronto-occipital fasciculus (IFOF), inferior longitudinal fasciculus (ILF), superior longitudinal fasciculus (SLF) and uncinate fasciculus (UF). Anatomical tract locations are shown in Figure 1. These 10 tracts were selected to give comprehensive coverage of major white matter tracts in the brain to explore their relationship with motor function in this population, as microstructural alterations have been observed in case children across widespread areas of white matter (Spencer et al., 2021).

**Figure 1:**
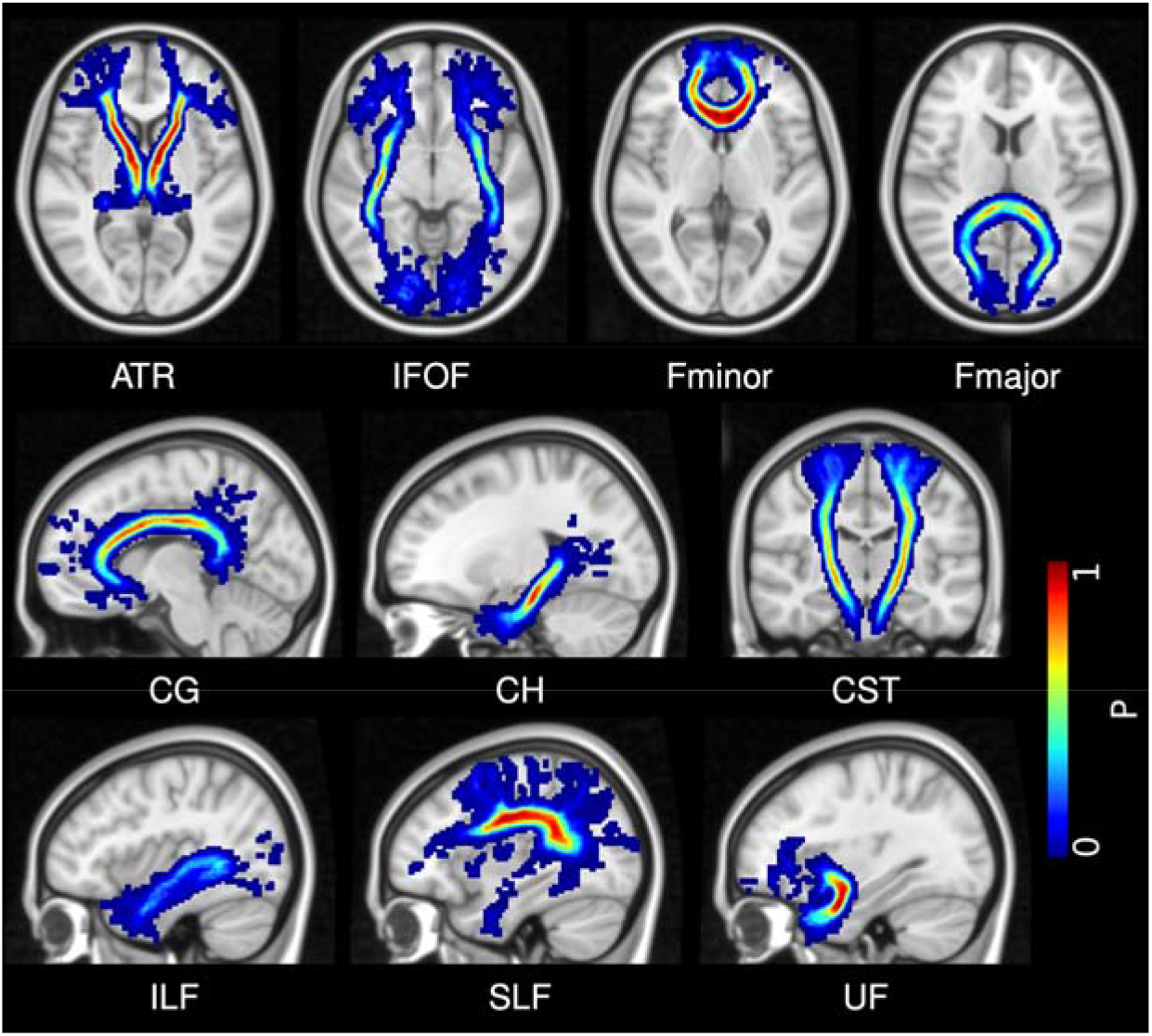
White matter tracts. We measured average FA in these 10 white matter tracts using an age-specific probabilistic white matter tract atlas (Spencer et al., 2020). The probability of a tract occupying any particular voxel is given by the colour bar. ATR = anterior thalamic radiation; IFOF = inferior fronto-occipital fasciculus; Fminor = forceps minor; Fmajor = forceps major; CG = cingulate gyrus part of the cingulum; CH = hippocampal part of the cingulum; CST = corticospinal tract; ILF = inferior longitudinal fasciculus; SLF = superior longitudinal fasciculus; UF = uncinate fasciculus.

To measure FA in each tract, FA images were generated by fitting a tensor model to the DWI data using the weighted least squares method in FSL’s FDT software. The average FA within each white matter tract was then measured using an age-specific probabilistic atlas of white matter tracts, which was constructed from the control group from this cohort as part of a separate study and has been shown to give better delineation of white matter tracts in this age group than an adult atlas (Spencer et al., 2020). Tract FA was calculated as the mean FA across all voxels within the atlas mask, with each voxel weighted by the probability given by the atlas mask.

### Structural Networks

We constructed a structural connectivity network for each subject (Figure 2). Nodes were defined by parcellating the T1-weighted image into 84 regions, as defined by the Desikan-Killiany atlas (Desikan et al., 2006), using FreeSurfer (http://surfer.nmr.mgh.harvard.edu) (Fischl, 2012). The FIRST (Patenaude et al., 2011) subcortical segmentation tool from FSL was combined with the cortical parcellation from FreeSurfer using the labelsgmfix tool from MRtrix 3.0 (www.mrtrix.org) (Tournier et al., 2019), as this gave better segmentation of subcortical structures than FreeSurfer (including the hippocampus and amygdala).

**Figure 2:**
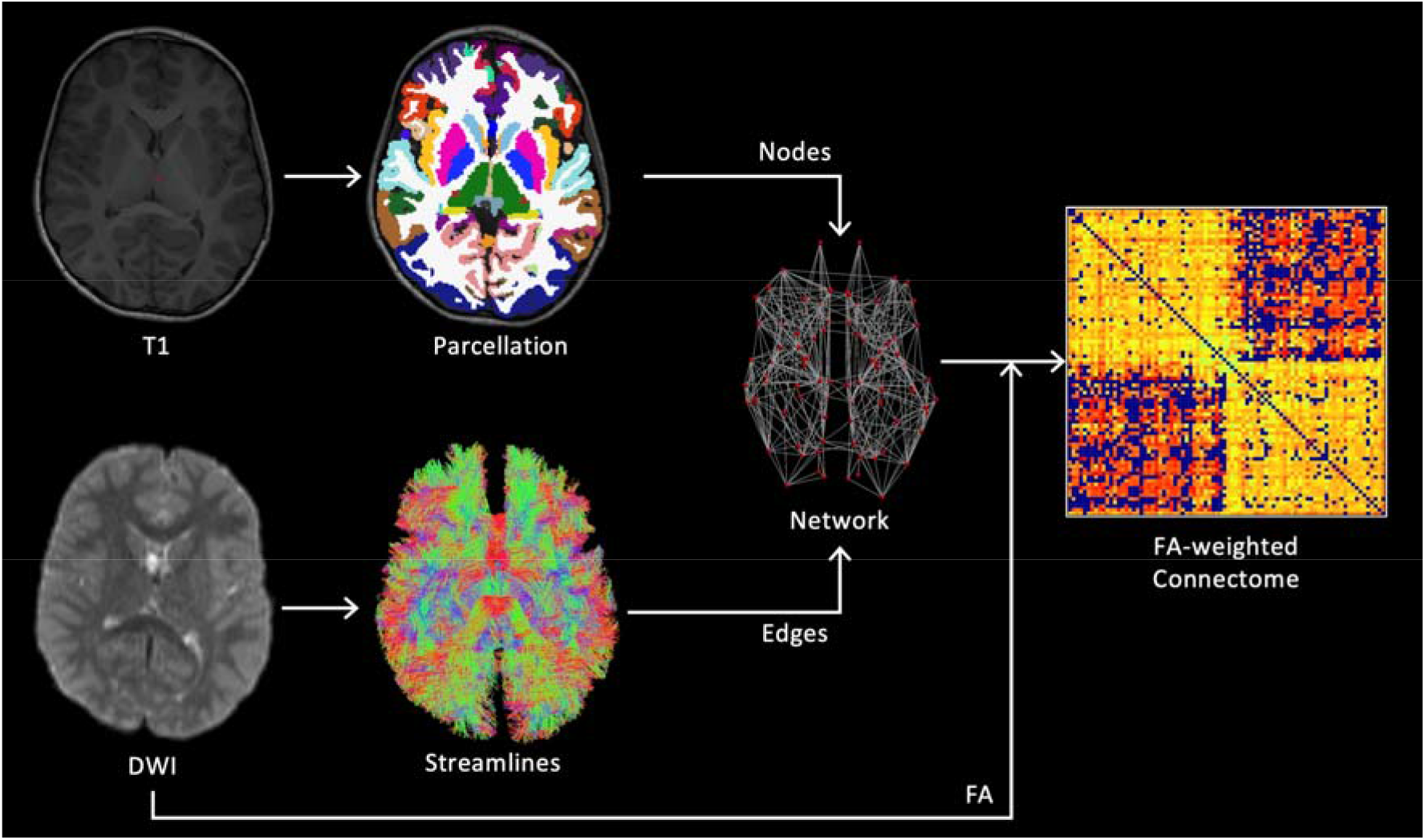
Method for constructing structural connectivity networks from T1 and DWI data. Nodes were defined by segmentation of the T1-weighted image. Edges were determined by probabilistic tractography and weighted by the mean FA along all streamlines connecting the corresponding pair of nodes. The resulting network was represented by a connectivity matrix. Reproduced from Spencer et al. (2021).

DWI processing and tractography steps, to define network edges, were performed using MRtrix. The DWI signal for a typical fibre population (the response function) was estimated from the data (Tournier et al., 2013) and used to calculate the fibre orientation distribution (FOD) by deconvolving the response function from the measured DWI signal using constrained-spherical deconvolution (Tournier et al., 2007). A five-tissue-type segmentation was generated from the T1-weighted image and used to perform anatomically-constrained tractography with the normalised FOD image (Smith et al., 2012). Tractography was performed using second-order integration over FODs (Tournier et al., 2010), with step size = 1 mm, minimum length = 50 mm, cutoff FOD magnitude = 0.1, and maximum angle between steps = 30°. Streamlines were seeded in the interface between grey and white matter and accepted if they terminated in subcortical or cortical grey matter (Smith et al., 2012). This method was used to generate 10 million streamlines which were subsequently filtered to 1 million using spherical-convolution informed filtering of tractograms (Smith et al., 2013) in order to improve biological plausibility and remove length bias. The weighted network for each subject was then constructed by defining an edge between any pair of nodes connected by at least one streamline, with edge weight defined by the mean FA along all streamlines connecting two nodes.

### Network Metrics

We measured graph-theoretic properties of the FA-weighted structural connectivity network using the following metrics: average strength, characteristic path length, global efficiency, local efficiency, clustering coefficient, modularity and small-worldness. For an in-depth description of these metrics, see Rubinov and Sporns (2010).

The node strength is the sum of the weights of all edges connected to a given node. The mean across all nodes gives the average strength for the network. The characteristic path length is the average shortest path between all pairs of nodes, where edge distances are defined inversely to edge weights, making stronger connections equivalent to shorter paths (note that this does not reflect physical distance between regions in the brain). The global efficiency is the average over all node pairs of the inverse of the shortest path length. The characteristic path length and global efficiency both reflect levels of global brain connectivity (with characteristic path length being more dependent on longer paths and global efficiency more dependent on shorter paths), and are both measures of the potential for integrated processing (Bullmore and Sporns, 2009; Rubinov and Sporns, 2010).

The local efficiency of a given node i is the inverse shortest path length between each pair of neighbours of i, averaged over all pairs of neighbours of i. This is then averaged across all nodes to give a measure for the whole network. The clustering coefficient measures the number of connections between the immediate neighbours of a node as the ratio of the number of actual edges between the immediate neighbours, modulated by edge weight, to the maximum possible number of such edges. The modularity indicates how well the network can be split up into relatively separate communities (i.e. modules) of nodes by measuring a normalised ratio of the number of within-module connections to the number of between-module connections, where the optimal modular structure is estimated with optimisation algorithms. The local efficiency, clustering coefficient and modularity indicate the efficiency of local information transfer, thus reflecting the potential for segregated functional processing (Bullmore and Sporns, 2009; Rubinov and Sporns, 2010).

Both integration and segregation are required for brain networks to carry out localised and distributed processing simultaneously (Tononi et al., 1994). The degree to which a network exhibits both segregation and integration is measured by the small-worldness of the network (Muldoon et al., 2016; Rubinov and Sporns, 2010). A high degree of small-worldness is characterised by a high clustering coefficient and low characteristic path length compared to random graphs. We measured small-worldness with small-world propensity (Muldoon et al., 2016). All other metrics were calculated with the Brain Connectivity Toolbox (http://www.brain-connectivity-toolbox.net) (Rubinov and Sporns, 2010).

### Statistical Analysis

We measured the partial Pearson correlation coefficient between tract FA and MABC-2 total score, with age and sex included as covariates, in the case and control group separately. We then measured the partial Pearson correlation coefficient between each network metric and MABC-2 total score, with age and sex included as covariates, in the case and control group separately. In each of the above tests, false discovery rate (FDR)-correction was applied to the two-tailed P-values using the Benjamini-Hochberg method (Benjamini and Hochberg, 1995). Results with FDR-corrected P<0.05 were considered significant. Correlation analysis was performed with MATLAB (R2019b, Mathworks).

To explore the relationship between connectivity and motor function, we used NBS (Zalesky et al., 2012, 2010) to test for group differences in the association between edge weights and MABC-2 total score. NBS is a nonparametric permutation-based approach for controlling family-wise error rate (FWER) on the level of subnetworks using the following procedure: 1) an F-test was performed on each edge to test for group differences in the slope between edge weight and MABC-2 total score; 2) edges were removed if the corresponding F-statistic was below a threshold of F_1,52_ = 7.1488 (equivalent to keeping edges with P<0.01); 3) of the remaining edges, the size (i.e. number of edges) of any connected subnetwork was stored; 4) this process was repeated for 10,000 random permutations of the data to estimate the null distribution; 5) the FWER-corrected P-value for a subnetwork was then calculated as the number of permutations for which the largest connected subnetwork in the permuted data was the same size or larger than the given subnetwork, normalised by the number of permutations. To further explore edge-level association with domains of the motor assessment, we repeated this analysis to test for group differences in the dependence of edge weight on each MABC-2 subscale (aiming and catching, balance, and manual dexterity). In order to only test robust edges, only connections present in >50% of children in each of the case and control group were assessed. Age and sex were included as covariates in a general linear model in all tests. Results with FWER-corrected P<0.05 were considered significant.

### Visualisation

Subnetworks resulting from NBS analysis were visualised with BrainNet Viewer (https://www.nitrc.org/projects/bnv/) (Xia et al., 2013) and as Circos connectograms (http://www.circos.ca) (Krzywinski et al., 2009).

## Results

### Participant Demographics

We recruited 51 case and 43 control children for this study (Figure 3). Of these, 7 cases and 4 controls did not want to undergo scanning, and an additional 4 cases had incomplete data due to movement during the scan. Of these 79 scans, DWI quality control led to the rejection of scans of 6 cases and 2 controls, and a further scan of one child from each group was rejected due to incorrect image volume placement. This left 33 case and 36 control scans which passed the DWI quality control and were used in the tract analysis. Of these, the T1-weighted anatomical scans for 11 cases and 4 controls were not of sufficient quality to allow segmentation and parcellation, leaving 22 cases and 32 controls for structural network analysis. Participant demographics are shown in Table 1.

**Table 1:** Participant demographics for each of the tract analysis and network analysis groups. Perinatal clinical information, as well as scores from neonatal MRI assessment of basal ganglia and thalami (BGT), white matter (WM) and posterior limbs of the internal capsule (PLIC), are given for cases. P-values are shown for case-control comparison of age, index of multiple deprivation, and MABC-2 scores using Wilcoxon rank sum tests, and for case-control comparison of sex, and MABC-2 scores <15^th^ centile using Fisher’s exact test. *P < 0.05. aEEG = amplitude integrated electroencephalogram; MABC-2 = Movement Assessment Battery for Children, Second Edition; TH = therapeutic hypothermia.

|  | Tract Analysis |  |  | Network Analysis |  |  |
| --- | --- | --- | --- | --- | --- | --- |
|  | Case (n = 33) | Control (n = 36) | p | Case (n = 22) | Control (n = 32) | p |
| Age, median (range) | 6.9 (6.0–7.9) | 7.0 (6.1–7.9) | 0.5555 | 7.0 (6.0–7.8) | 7.0 (6.1–7.8) | 0.5428 |
| Sex, male/female | 18/15 | 19/17 | 0.8894 | 12/10 | 16/16 | 0.7526 |
| Index of Multiple Deprivation, median (range) | 7 (1–10) | 7 (2–10) | 0.5211 | 7 (1–10) | 7 (2–10) | 0.8174 |
| **MABC-2 Assessment:** |  |  |  |  |  |  |
| MABC-2 total score, median (range) | 9 (3-19) | 11 (5-18) | 0.0866 | 10 (3-19) | 11 (5-18) | 0.3955 |
| MABC-2 total score <15<sup>th</sup> centile, n (%) | 12 (36) | 2 (6) | 0.0021* | 9 (41) | 2 (6) | 0.0041* |
| Aiming & catching score, median (range) | 9 (2-15) | 9 (5-18) | 0.5090 | 10 (4-15) | 9 (5-18) | 0.6967 |
| Aiming & catching score <15<sup>th</sup> centile, n (%) | 6 (18) | 2 (6) | 0.1401 | 4 (18) | 2 (6) | 0.2111 |
| Balance score, median (range) | 9 (3-19) | 11 (5-15) | 0.0782 | 10 (4-19) | 11.5 (5-15) | 0.4187 |
| Balance score <15<sup>th</sup> centile, n (%) | 7 (21) | 3 (8) | 0.1767 | 3 (14) | 2 (6) | 0.3875 |
| Manual dexterity score, median (range) | 9 (1-18) | 11 (3-18) | 0.0773 | 9 (1-18) | 11 (3-18) | 0.2620 |
| Manual dexterity score <15<sup>th</sup> centile, n (%) | 11 (33) | 4 (11) | 0.0397* | 7 (32) | 4 (13) | 0.0996 |
| **Neonatal MRI Assessment:** |  |  |  |  |  |  |
| BGT, median (range) | 0 (0–2) | - | - | 0 (0–1) | - | - |
| WM, median (range) | 1 (0–3) | - | - | 1 (0–3) | - | - |
| PLIC, median (range) | 0 (0–2) | - | - | 0 (0–1) | - | - |
| **Perinatal Clinical Information:** |  |  |  |  |  |  |
| Mode of delivery, vaginal/instrumental/emergency caesarean in labour/emergency caesarean not in labour | 11/8/10/4 | - | - | 8/6/7/1 | - | - |
| Assisted ventilation at 10 minutes of age, yes/no | 24/9 | - | - | 17/5 | - | - |
| Cardiac compressions required, yes/no | 13/20 | - | - | 8/14 | - | - |
| Apgar score at 10 minutes of age, median (range) | 6 (0–10) | - | - | 5 (0–10) | - | - |
| Worst pH within 1 hour of birth, median (range) | 6.98 (6.70–7.25) | - | - | 6.98 (6.77–7.25) | - | - |
| Worst base excess within 1 hour of birth, median (range) | -16.0 (-31.0 to -4.8) | - | - | -16.1 (-31.0 to -4.8) | - | - |
| Grade of encephalopathy: moderate, n (%) | 25 (76) | - | - | 18 (82) | - | - |
| Grade of encephalopathy: severe, n (%) | 8 (24) | - | - | 4 (18) | - | - |
| aEEG abnormalities prior to TH: moderate, n (%) | 30 (91) | - | - | 20 (91) | - | - |
| aEEG abnormalities prior to TH: severe, n (%) | 3 (9) | - | - | 2 (9) | - | - |

**Figure 3:**
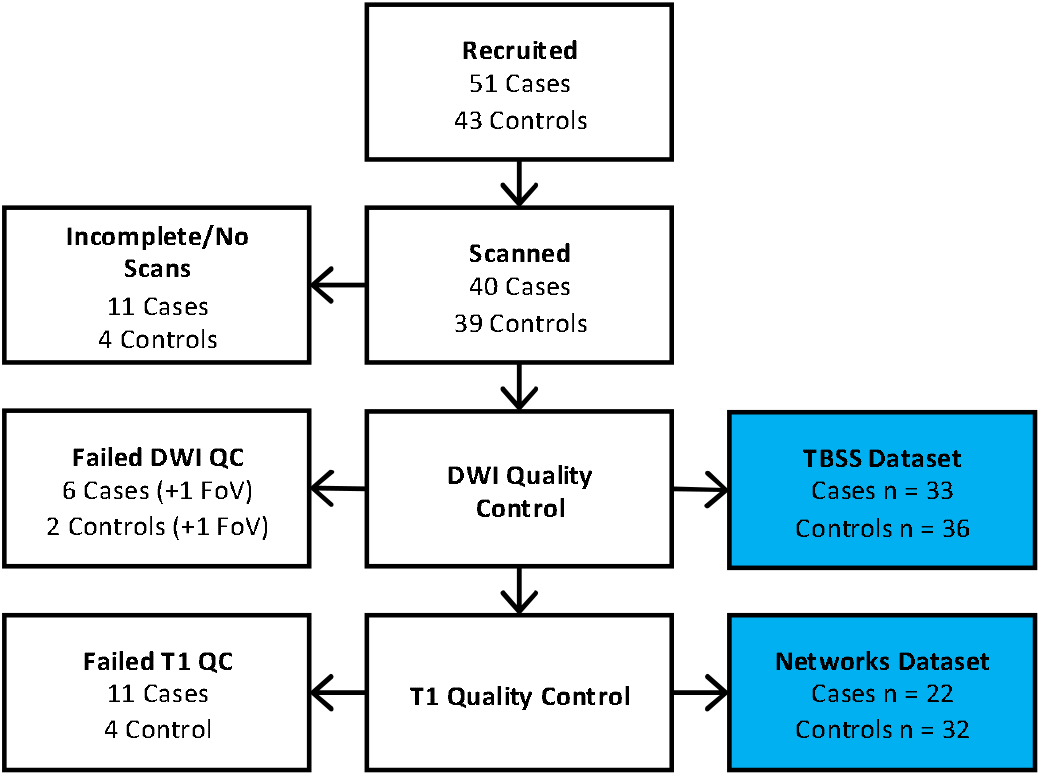
Flowchart of participants at each stage of recruitment and quality control. FoV = field of view, indicating the scans which were rejected due to incorrect image volume placement.

Anatomical images were visually assessed for focal lesions and abnormal signal intensities. Small, non-specific white matter signal changes were seen in 1 case and 2 controls in the tract analysis dataset, and in 1 control in the network analysis dataset. These findings were judged by a blinded assessor (FC) to be minor and the scans were therefore not excluded.

Perinatal clinical information and scores from qualitative assessment of neonatal MRI is also shown in Table 1. Median scores from assessment of neonatal MRI were low, as expected in this cohort and the findings unlikely to lead to motor problems from current data and 2-year follow-up. When measured with Kendall’s Tau rank correlation, there was no association between MABC-2 score and neonatal MRI scores for basal ganglia and thalami (r = -0.181, p = 0.2272), white matter (r = -0.202, p = 0.1509), or posterior limb of the internal capsule (r = -0.226, p = 0.130).

Table 1 also shows that, similar to previously reported findings from a subset of participants from this cohort (Lee-Kelland et al., 2020), a significantly higher proportion of case children had MABC-2 total scores less than the 15^th^ centile than controls in both the tract analysis cohort (P = 0.0021) and the network analysis cohort (P = 0.0041). Additionally, a higher proportion of case children had manual dexterity scores less than the 15^th^ centile than controls in the tract analysis cohort (P = 0.0397), but not the network analysis cohort (P = 0.0996).

### Tract Microstructure

We measured the partial correlation between tract-level FA and MABC-2 total score. Significant correlations were found in the case group with the ATR bilaterally (left: r = 0.513, P = 0.0191; right: r = 0.488, P = 0.0191), left CG (r = 0.588, P = 0.0090), left CH (r = 0.541, P = 0.0153), and IFOF bilaterally (left: r = 0.445, P = 0.0363; right: r = 0.494, P = 0.0191). In the control group, the relationship between tract-level FA and MABC-2 total score exhibited the same trends as in the case group, but no correlations were significant. Tracts which exhibited significant correlations are plotted for both groups in Figure 4. Correlation coefficients for all tracts, as well as uncorrected P-values, are given in Supplementary Table 1.

**Figure 4:**
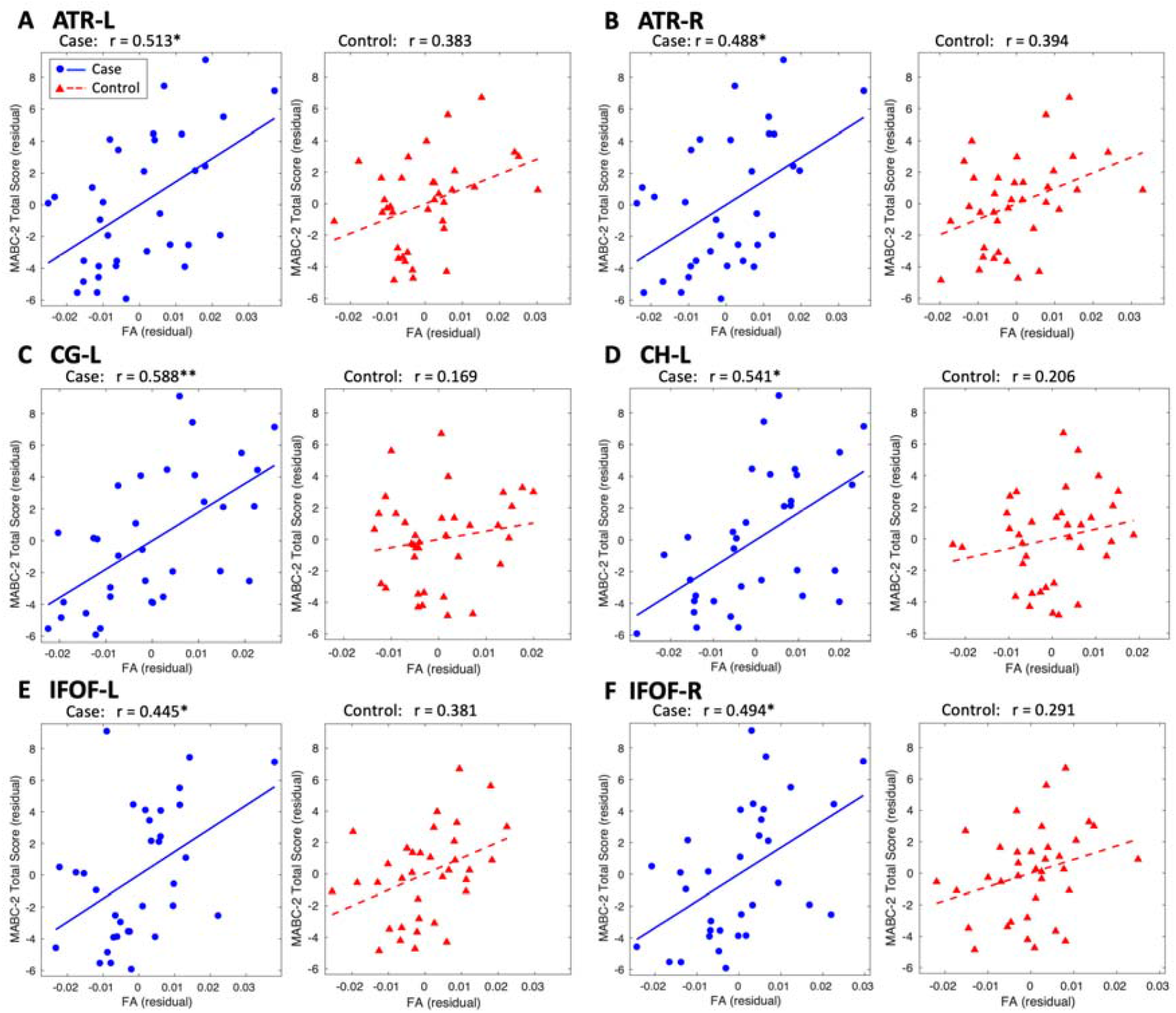
Correlation of tract FA with MABC-2 total score. The partial Pearson correlation was measured between tract-level FA and MABC-2 total score, controlled for age and sex. Significant results were only found in cases. For these results, residuals are plotted for cases (blue circles, solid blue line), with controls also shown for comparison (red triangles, dashed red line). FDR-corrected: *P<0.05, **P<0.01.

### Structural Network Metrics

We measured the partial correlation between structural network metrics and MABC-2 total score. In cases, significant correlations were found with average node strength, local efficiency, global efficiency, clustering coefficient, and characteristic path length. In the control children, the relationship between network metrics and MABC-2 total score exhibited the same trend as in the case children, but no correlations were significant. All metrics are plotted for both cohorts in Figure 5.

**Figure 5:**
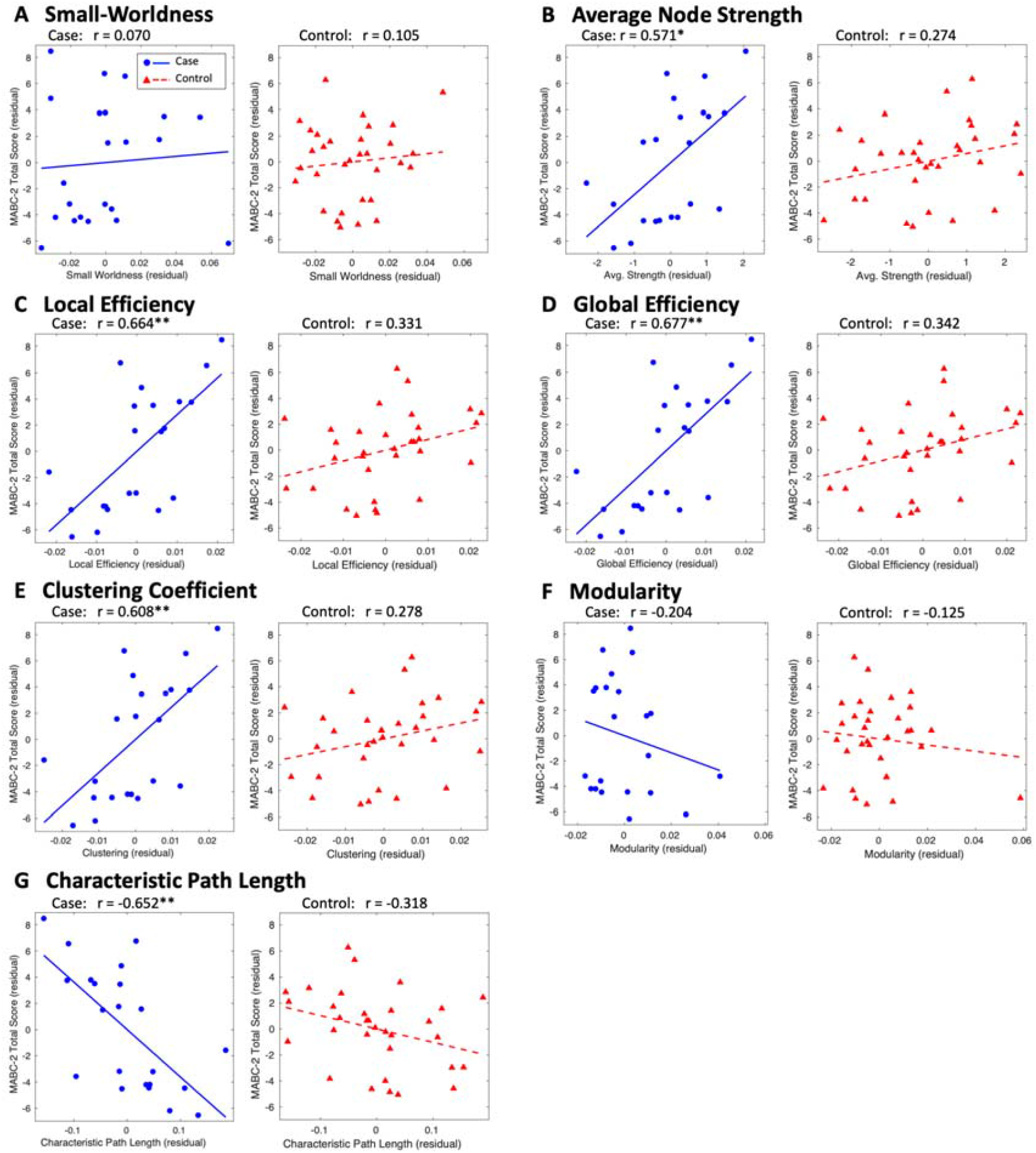
Correlation of structural network metrics with MABC-2 total score. The partial Pearson correlation was measured between structural network metrics and MABC-2 total score, controlled for age and sex. Residuals are plotted for cases (blue circles, solid blue line) and controls (red triangles, dashed red line). FDR-corrected: *P<0.05, **P<0.01.

### Network-Based Statistic

Group differences were found in the dependence of MABC-2 total score on edge weight (P = 0.0109, Figure 6) in a subnetwork comprising 24 nodes (18 left, 6 right) and 34 edges (13 interhemispheric, 21 intrahemispheric in the left hemisphere). The nodes in the subnetwork with the most connections include the rostral middle frontal gyrus bilaterally, superior frontal gyrus bilaterally, left insula, left pars opercularis, left caudal anterior cingulate, pars triangularis bilaterally, putamen bilaterally. Also included in the subnetwork were the left inferior temporal gyrus, left middle temporal gyrus, rostral anterior cingulate gyrus bilaterally, left superior temporal gyrus, left supramarginal gyrus, left temporal pole, left cerebellum, left banks of the superior temporal sulcus, left caudal middle frontal gyrus, left inferior parietal gyrus, left precentral gyrus, and right accumbens area.

**Figure 6:**
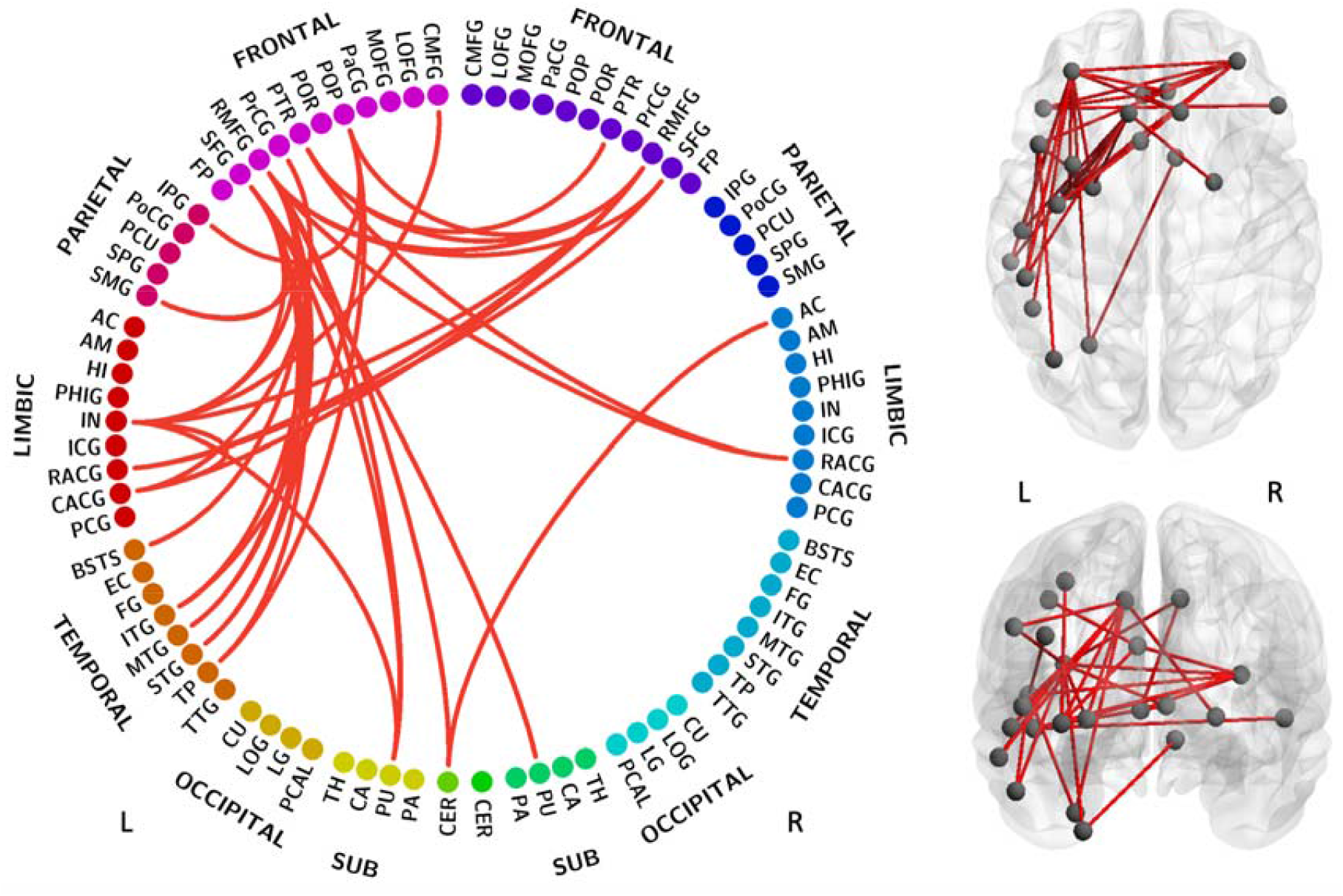
MABC-2 total score subnetwork. The subnetwork shown exhibited group differences in the dependence of MABC-2 total score on edge weights (P = 0.0109). Node label abbreviations are given in Supplementary Table 3.

When exploring each subscale of the MABC-2 assessment, group differences were found in the association between balance scores and edge weight (P = 0.0245), and between manual dexterity scores and edge weight (P = 0.0233) but not between aiming and catching scores and edge weight (P > 0.05). The balance subnetwork (Figure 7A) comprised 17 nodes (10 left, 7 right) and 17 edges (9 interhemispheric, 4 intrahemispheric in the left hemisphere, 4 intrahemispheric in the right hemisphere). The nodes in the subnetwork with the most connections include the rostral middle frontal gyrus bilaterally, pars triangularis bilaterally, and insula bilaterally. The manual dexterity subnetwork (Figure 7B) comprised 21 nodes (15 left, 6 right) and 21 edges (5 interhemispheric, 14 intrahemispheric in the left hemisphere, 2 intrahemispheric in the right hemisphere). The nodes in this subnetwork with the most connections include the left superior frontal gyrus, insula bilaterally, left rostral middle frontal gyrus, left putamen, and right caudate. The complete list of nodes in each subnetwork is given in Supplementary Table 2.

**Figure 7:**
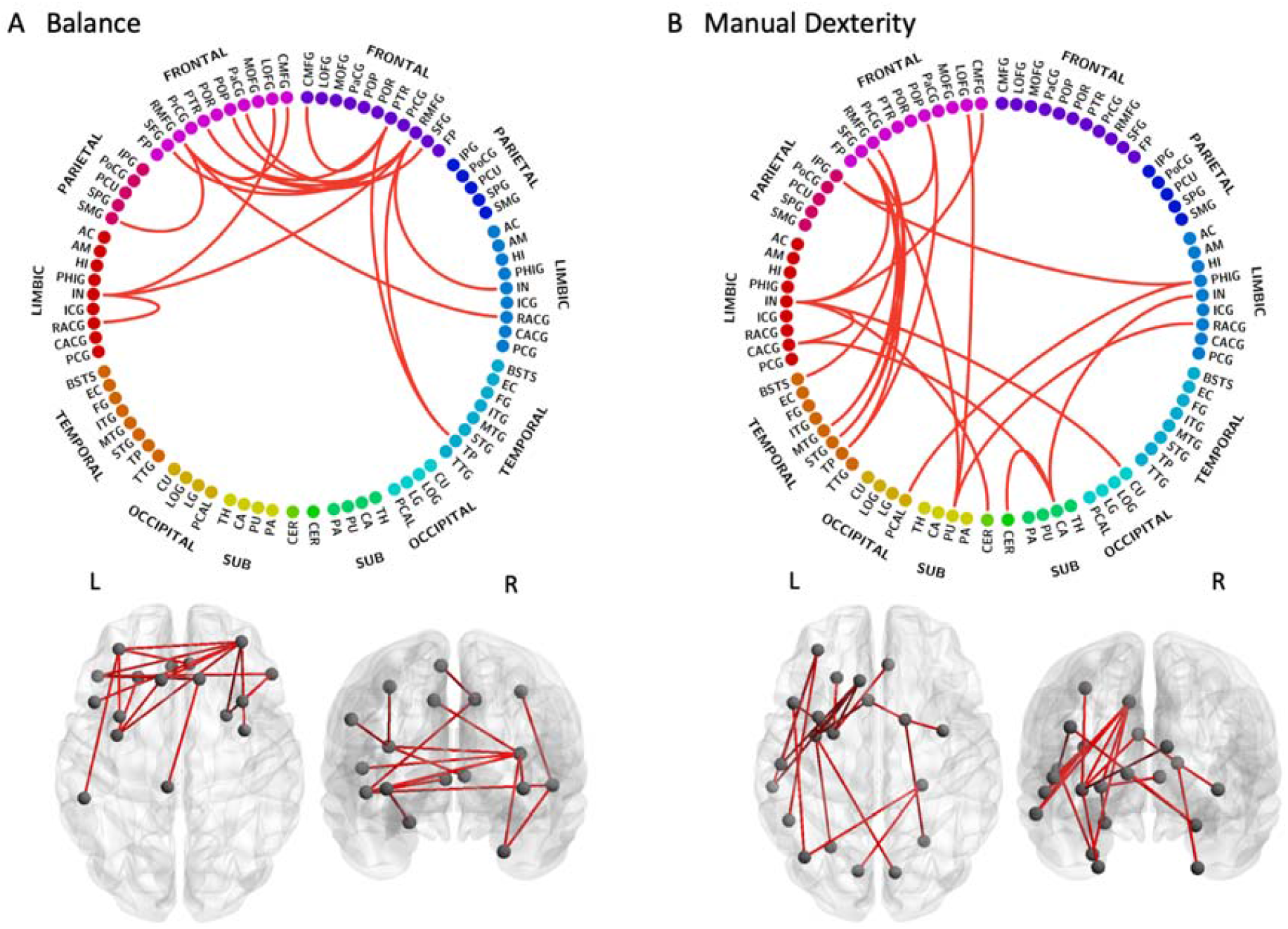
Balance and manual dexterity subnetworks. The subnetworks shown exhibit group differences in the dependence of MABC-2 subscale scores on edge weight for: A) balance (P = 0.0245); and B) manual dexterity (P = 0.0233). Node label abbreviations are given in Supplementary Table 3.

## Discussion

In this study, we investigated the association between motor performance and white matter connectivity in a cohort of early school-age children without CP, who were treated with TH for HIE at birth, and controls matched for age, sex and socio-economic status. Assessment of tract-level white matter microstructure revealed an association between MABC-2 total score and FA in the ATR bilaterally, left CG, left CH, and IFOF bilaterally in cases. No such relationship was observed in controls. We constructed a structural connectivity network for each subject to investigate the association between whole-brain connectivity properties and motor function. We found correlations between MABC-2 total score and average node strength, local efficiency, global efficiency, clustering coefficient, and characteristic path length in cases. As before, no significant correlations were found in controls. We then investigated edge-level associations between connectivity and MABC-2 total score, as well as each of the MABC-2 subscale scores (aiming and catching, balance, and manual dexterity). This revealed subnetworks, in which the association between motor outcome and edge weight was significantly different between cases and controls, for MABC-2 total scores as well as balance and manual dexterity subscale scores.

### Tract Microstructure Correlates with Motor Function

Studies on children with NE, prior to widespread use of TH, have shown that the presence of lesions on neonatal MRI are strongly associated with both early motor deficits and the absence of lesions is strongly associated with no motor deficit at 2-3 years (Martinez-Biarge et al., 2012; Rutherford et al., 1996) and school-age motor outcome of children without CP (van Kooij et al., 2010). In infants treated with TH (including those who go on to develop CP), neonatal MRI was predictive of early disability (Rutherford et al., 2010), and FA in the corticospinal tract and anterior centrum semiovale on neonatal MRI was associated with motor function at around 2 years of age (Massaro et al., 2015; Tusor et al., 2012). In cooled infants without CP, motor function at 2 years of age correlated with FA in the posterior limb of the internal capsule, centrum semiovale, corpus callosum, left cerebral peduncle and brain stem (Tusor et al., 2012). In this study we have shown, in cooled infants without CP at early school age, that the increased risk of motor impairments (Jary et al., 2019; Lee-Kelland et al., 2020) is associated with microstructural differences in the ATR bilaterally, left CG, left CH, and IFOF bilaterally. The absence of significant correlations in the control group, despite these tracts being associated with motor function in case children, suggests a ceiling effect whereby the motor performance of control children is more dependent on other factors, whereas the restricted diffusion in these tracts in cases is associated with impaired motor function.

A few of these children had minor basal ganglia, thalamic and white matter lesions on their neonatal scans, which may have impaired the development of white matter tracts. However, median scores were low, thus not expected to result in motor problems (Rutherford et al., 2010), and lesions were not sufficient to be evidenced by overt abnormality on the later scans in the vast majority. The neonatal MRI scoring system may be better for predicting gross motor deficits observable at an earlier age, than the more subtle impairments assessed through MABC-2 that only become accessible at a later age.

### Whole-Brain Connectivity Correlates with Motor Outcome

We previously reported no significant differences in group means of network metrics between cases and controls (Spencer et al., 2021). Despite this, and despite finding no correlation between network metrics and motor ability in controls, we found a close relationship between network metrics and motor ability in cases. Thus, white matter connectivity and brain organisation are more predictive of motor function in case children than in controls.

A large local efficiency and a large clustering coefficient are both indicators of the potential for functional segregation (i.e. carrying out localised processing amongst locally connected brain regions) (Rubinov and Sporns, 2010). Conversely, a large global efficiency and a small characteristic path length are both indicators of the potential for functional integration (i.e. carrying out distributed processing, combining information from distant brain areas) (Bullmore and Sporns, 2009; Rubinov and Sporns, 2010). In cases, we found trends of increasing local efficiency and clustering coefficient with increased motor ability, in addition to increasing global efficiency and decreasing characteristic path length with increased motor ability, indicating that case children with impaired motor function have reduced levels of brain network segregation and integration. This trend has previously been observed in a cohort of 6-month-old infants exposed to HIE (Tymofiyeva et al., 2012). In the developing brain, alterations to network segregation and integration are thought to be associated with pruning and myelination respectively (Dennis and Thompson, 2013a; Tymofiyeva et al., 2014). The impact of hypoxic injury on these mechanisms (Gressens et al., 2008; O’Brien et al., 2019) may cause long-term alterations to white matter development (Hüppi and Dubois, 2006) which result in the altered dependence of motor function on connectivity in case children at school age.

### Edge-Level Connectivity Correlates with Motor Outcome

NBS analysis revealed subnetworks which expressed group differences in the association between edge weight and motor function for MABC-2 total score as well as balance and manual dexterity. No significant result was found for aiming and catching score, possibly due to the small sample size. The MABC-2 total score subnetwork was left-lateralised, with numerous fronto-temporal connections as well as some connections to limbic structures. Frontal lobe regions in the MABC-2 total score subnetwork with known motor association include the superior frontal gyrus, and rostral and caudal middle frontal gyri. The superior frontal gyrus contains the supplementary motor area (Martino et al., 2011; Nachev et al., 2008) and makes long-range connections to the parietal, occipital and temporal lobes via the IFOF and cingulum (Briggs et al., 2020). The rostral and caudal middle frontal gyri, as defined by the Desikan-Killiany atlas, correspond to the dorsolateral prefrontal cortex and the premotor area, respectively (Desikan et al., 2006; Kikinis et al., 2010). The premotor area is involved in movement representation (Rizzolatti et al., 1996). The dorsolateral prefrontal cortex is involved in higher-level cognitive functions including working memory (Barbey et al., 2013) and attention (Japee et al., 2015), but is also associated with motor function and makes connections to the premotor area, supplementary motor area and cerebellum (Diamond, 2000).

Additional regions in the MABC-2 subnetwork with known involvement in motor function include the putamen (movement regulation and sensorimotor coordination), cerebellum (motor control), precentral gyrus (primary motor area), inferior parietal gyrus (motor function and action representation (Fogassi and Luppino, 2005)), and caudal anterior cingulate gyrus (involved in motor control and is connected to the primary and supplementary motor areas (Koski and Paus, 2000; Stevens et al., 2011)). The MABC-2 subnetwork also included several regions involved in visual processing, including the superior temporal gyrus (visual information integration (Karnath, 2001)), inferior temporal gyrus (visual processing and visual object recognition (Conway, 2018)), and the banks of the superior temporal sulcus (visual attention and goal-direction action (Shultz et al., 2011)).

In addition to these brain regions with known involvement in motor function and visual processing, the MABC-2 subnetwork comprised several areas associated with higher-level cognitive function, including the rostral anterior cingulate gyrus (emotion and cognition (Stevens et al., 2011)), insula (sensorimotor processing as well as emotion, attention and salience processing (Uddin et al., 2017)), and the pars opercularis and pars triangularis, which are both involved in language but are also associated with in motor function (Tamada et al., 1999). The inclusion of these areas possibly reflects the involvement of cognitive processes in motor function or in understanding and attending to the tasks within the MABC-2 assessment, or may be due to the association between motor impairment and reduced cognitive abilities in case children (Jary et al., 2019).

The majority of the connections in the balance subnetwork were between areas in the frontal lobes, with the addition of some limbic areas. Many regions in the balance subnetwork were also present in the MABC-2 total score subnetwork and are associated with motor function, including the rostral and caudal middle frontal gyri, superior frontal gyrus, insula, pars opercularis and pars triangularis. The balance subnetwork also included the paracentral gyrus which is involved in sensorimotor processing.

The manual dexterity subnetwork comprised several fronto-temporal connections in the left hemisphere, in addition to connections to limbic and subcortical areas. Many of these regions are involved in motor function and were present in the MABC-2 total score subnetwork (rostral and caudal middle frontal gyri, superior frontal gyrus, caudal anterior cingulate gyrus, insula, pars opercularis, inferior parietal gyrus, putamen and cerebellum) with the addition of the caudate, which is involved in goal-directed action and cognition. The manual dexterity subnetwork also included several areas involved in visual processing, for example the superior temporal gyrus and banks of the superior temporal sulcus, which were also present in the MABC-2 subnetwork, as well as the pericalcarine cortex (visual association area) and cuneus cortex (basic visual processing). The inclusion of visual processing areas in this subnetwork, and in the MABC-2 subnetwork, possibly reflects the visual processing demands of constituent tasks in the MABC-2 assessment.

The MABC-2 total score subnetwork and the manual dexterity subnetwork both contained many intrahemispheric connections in the left hemisphere. Additionally, tract-level analysis revealed correlations in cases between MABC-2 total score and FA in the left CG and CH but not right. Conversely, in our previous work we found a right-lateralised subnetwork which exhibited group differences in the dependence of edge weight on processing speed (Spencer et al., 2021), despite finding no laterality in the areas of white matter with reduced FA in cases. Additionally, laterality of FA alterations has not been reported in neonates treated with TH (Tusor et al., 2012). Therefore, the laterality observed in relation to functional outcome at school-age in cases is unlikely to be due to targeted injury – injury patterns following HIE are generally bilateral and fairly symmetrical.

All case children included in the network analysis dataset were right-handed, however the handedness data for the rest of the cohort were incomplete. In a very large population study of white matter in typically developing school-age children, no relationship was found between handedness and white matter microstructure after correcting for multiple comparisons (López-Vicente et al., 2021). The laterality observed in relation to motor outcome in cases may be the result of compensatory mechanisms causing structural alterations that are differentially highlighted by different domains of the motor assessments. These mechanisms may affect one hemisphere more than the other depending on the individual’s handedness, and become apparent when observed in comparison with controls due to the lack of association between handedness and microstructure in typically developing children (López-Vicente et al., 2021).

### Strengths and Limitations

To our knowledge, this is the first study to explore the relationship between brain structural connectivity and motor function in school-age children treated with TH for NE, who did not develop CP. Anatomically-constrained tractography was performed on high angular resolution DWI data, using a method capable of resolving crossing fibres (Tournier et al., 2012, 2008). For tract-level analysis, we used a probabilistic atlas of white matter tracts constructed from the control group, which gives better delineation of white matter tracts in this age group than an adult atlas (Spencer et al., 2020). Movement artefacts are common in data acquired from paediatric populations (Phan et al., 2018). Therefore, we used a thorough quality control pipeline to reject poor quality scans. This resulted in a relatively small sample size, which is a possible limitation of this study. To ensure robustness of the NBS results, we only tested edges which were non-zero in the connectomes of over half the subjects in both the case and control groups. The handedness data for the cohort were incomplete, therefore we were unable to quantitatively investigate the involvement of handedness in the association between motor function and white matter microstructure. Further study is needed to explore the involvement of handedness in the development of white matter and motor function in case children.

## Conclusion

We have identified edge-level, tract-level and whole-brain structural connectivity properties which correlate with motor outcome assessed robustly in school-age children treated with TH for HIE at birth, who did not develop CP, but not in controls. Future longitudinal investigation of brain development from the neonatal period to school age would reveal how the early microstructural alterations, resulting from injury at birth, lead to the emergence of these correlations. This would inform therapeutic strategies to promote healthy development of motor function.

## Data Availability

The raw data that support these findings are available upon reasonable request to the corresponding author.

## Acknowledgements

We thank the children and their families for participating, Ngoc Jade Thai for her assistance with MR sequences and Aileen Wilson for her radiographical expertise.

