## Supplementary Materials for "Motor Function and White Matter Connectivity in Children Treated with Therapeutic Hypothermia for Neonatal Encephalopathy"

|  | Exposed cohort | | | Comparator cohort | | |
| --- | --- | --- | --- | --- | --- | --- |
| Tract | r | Uncorrected p | FDR-corrected p | r | Uncorrected p | FDR-corrected p |
| L ATR | 0.513 | 0.0032 | 0.0191 | 0.383 | 0.0255 | - |
| R ATR | 0.488 | 0.0053 | 0.0191 | 0.394 | 0.0213 | - |
| L CG | 0.588 | 0.0005 | 0.0090 | 0.169 | 0.3404 | - |
| R CG | 0.382 | 0.0338 | - | 0.002 | 0.9904 | - |
| L CH | 0.541 | 0.0017 | 0.0153 | 0.206 | 0.2434 | - |
| R CH | 0.395 | 0.0277 | - | 0.087 | 0.6262 | - |
| L CST | 0.228 | 0.2182 | - | 0.178 | 0.3146 | - |
| R CST | 0.140 | 0.4529 | - | 0.200 | 0.2566 | - |
| Fminor | 0.354 | 0.0506 | - | 0.402 | 0.0184 | - |
| Fmajor | 0.154 | 0.4095 | - | 0.215 | 0.2229 | - |
| L IFOF | 0.445 | 0.0121 | 0.0363 | 0.381 | 0.0261 | - |
| R IFOF | 0.494 | 0.0047 | 0.0191 | 0.291 | 0.0954 | - |
| L ILF | 0.208 | 0.2621 | - | 0.277 | 0.1128 | - |
| R ILF | 0.188 | 0.3123 | - | 0.105 | 0.5538 | - |
| L SLF | 0.352 | 0.0522 | - | 0.270 | 0.1223 | - |
| R SLF | 0.333 | 0.0671 | - | 0.337 | 0.0538 | - |
| L UF | 0.314 | 0.0851 | - | 0.248 | 0.1581 | - |
| R UF | 0.362 | 0.0455 | - | 0.020 | 0.9112 | - |

Supplementary Table 1: Tract-level correlations. Partial Pearson correlation coefficients, r, were measured for the exposed cohort and comparator cohort separately. All uncorrected p-values are shown, as well as all significant FDR-corrected p-values. Abbreviations: L = left hemisphere; R = right hemisphere; ATR = anterior thalamic radiation; CG = cingulate gyrus part of the cingulum; CH = hippocampal part of the cingulum; CST = corticospinal tract; Fminor = forceps minor; Fmajor = forceps major; IFOF = inferior fronto-occipital fasciculus; ILF = inferior longitudinal fasciculus; SLF = superior longitudinal fasciculus; UF = uncinate fasciculus.

| **MABC** | **Balance** | **Manual Dexterity** |
| --- | --- | --- |
| L & R RMFG | L & R RMFG | L SFG |
| L & R SFG | L & R PTR | L & R IN |
| L IN | L & R IN | L RMFG |
| L POP | L & R SFG | L PU |
| L CACG | R TP | R CA |
| L & R PTR | L & R CMFG | L CACG |
| L & R PU | L & R RACG | L IPG |
| L ITG | L LOFG | L STG |
| L MTG | L POP | L MTG |
| L & R RACG | L PaCG | L POP |
| L STG | L SMG | L TP |
| L SMG |  | R PHIG |
| L TP |  | L BSTS |
| L CER |  | L CMFG |
| L BSTS |  | L LOFG |
| L CMFG |  | L PCAL |
| L IPG |  | L & R CER |
| L PrCG |  | R CU |
| R AC |  | R RACG |

Supplementary Table 2: Complete list of nodes in each subnetwork. The node label abbreviation is preceded by L or R to indicate left or right hemisphere, respectively. Node label abbreviations are shown in Supplementary Table 3.

| ***Frontal*** |  |  | ***Limbic*** |  |
| --- | --- | --- | --- | --- |
| CMFG | Caudal middle frontal gyrus |  | AC | Accumbens area |
| FP | Frontal pole |  | AM | Amygdala |
| LOFG | Lateral orbital frontal gyrus |  | CACG | Caudal anterior cingulate gyrus |
| MOFG | Medial orbital frontal gyrus |  | HI | Hippocampus |
| PaCG | Paracentral gyrus |  | ICG | Isthmus of the cingulate gyrus |
| POP | Pars opercularis |  | IN | Insula |
| POR | Pars orbitalis |  | PCG | Posterior cingulate gyrus |
| PTR | Pars triangularis |  | PHIG | Parahippocampal gyrus |
| PrCG | Precentral gyrus |  | RACG | Rostral anterior cingulate gyrus |
| RMFG | Rostral middle frontal gyrus |  |  |  |
| SFG | Superior frontal gyrus |  |  |  |
| ***Parietal*** |  |  | ***Occipital*** |  |
| IPG | Inferior temporal gyrus |  | CU | Cuneus cortex |
| PCU | Precuneus cortex |  | LG | Lingual gyrus |
| PoCG | Postcentral gyrus |  | LOG | Lateral occipital gyrus |
| SMG | Supramarginal gyrus |  | PCAL | Pericalcarine cortex |
| SPG | Superior parietal gyrus |  |  |  |
| ***Temporal*** |  |  | ***Subcortical*** |  |
| BSTS | Banks of the superior temporal |  | CA | Caudate |
|  | sulcus |  | PA | Pallidum |
| EC | Entorhinal cortex |  | PU | Putamen |
| FG | Fusiform gyrus |  | TH | Thalamus |
| ITG | Inferior temporal gyrus |  |  |  |
| MTG | Middle temporal gyrus |  | CER | Cerebellum |
| STG | Superior temporal gyrus |  |  |  |
| TP | Temporal pole |  |  |  |
| TTG | Tranverse temporal gyrus |  |  |  |

Supplementary Table 3: Node label abbreviations.
